# Service Retention Among Coast Guard Members Seeking Behavioral Healthcare

**DOI:** 10.1101/2023.07.19.23292893

**Authors:** John Allen, Mary Vance, Jerry Mahlau-Heinert, Jaspal Ahluwalia, Dana Thomas, Swati Singh, John Iskander

## Abstract

**Introduction:** Behavioral health conditions (BHC) can reduce service member retention. This analysis sought to identify demographic and diagnostic factors among BHC care-seeking Active-Duty United States Coast Guard (ADCG) that were predictive of discharge before completion of obligated service.

**Methods:** A four-year retrospective cohort study of ADCG personnel was conducted. Five machine-learning (ML) algorithms and logistic regression were applied to data on ADCG who sought outpatient care for BHC in 2016. Covariates examined as possible mediators of early service termination included diagnosis group, gender, rank grouping, and race.

**Results:** Only 26.4 of every 1,000 members who sought BHC care did not complete their service obligation. Diagnosis group did not predict early service termination, whereas senior enlisted rank was associated with early termination. The ML algorithms best predictive of early discharge from service were bagging classifier and decision tree classifier. Logistic regression performed as well as the two leading algorithms.

**Conclusions:** Specific ML models can be used to identify personnel groups at risk for early separation, such as senior enlisted personnel. Traditional epidemiologic methods demonstrate value in predicting service member separation.

## Background

Behavioral health conditions (BHC) can affect fitness for duty and service member retention. Understanding common risk factors that are predictive of Active-Duty Coast Guard service members (ADCG) not completing their service obligation is conducive to creating policies and initiatives to improve service member retention. This hypothesis-generating analysis sought to identify demographic factors and BHC diagnosis groups among ADCG that were predictive of discharge before completion of obligated service, an understudied area of inquiry within the Department of Defense (DoD). The general form of this study is survival analysis. Machine learning (ML) methods have been applied successfully to survival analysis both generally^1^ and specific to health related outcomes^2,3^. Multiple machine learning (ML) algorithms and a comparator analytic epidemiologic method were applied to data on ADCG who sought outpatient behavioral healthcare in 2016.

## Methods

Two source data sets were used; one was generated from the Health System Management Analysis and Reporting Tool (M2) database. M2 is a web-based query tool that provides data on health services provided to eligible beneficiaries in DoD military treatment facilities (MTFs) and private sector facilities. The second data set was generated from the Coast Guard human resources database, Personnel Services Command (PSC) Direct Access. A merged data set was created which included the M2 and PSC data sets: it contained information for ADCG who had outpatient visits to MTFs and/or private sector facilities from January 1 2016 to December 31 2019 (the study period), where the visits were under the Mental Health Product Line (i.e. care was received from mental and behavioral health specialists), and included BHC-related ICD10 codes used by the Defense Health Agency^4^. Encounters with non-mental health specialists were therefore not included in this analysis. New accessions were included if they were officer candidates (i.e., cadets), but enlisted recruits were excluded due to their high drop-out rate prior to completing basic training.

The merged final data set contained as independent variables rank group, most common (modal) primary BHC diagnosis group, race, gender, gender/race relative risk, unit type, number of visits, and time in service. Modal primary BHC diagnosis group was chosen to better represent the natural history of presenting illness over the course of the study. The data types and definitions for these variables are provided in Supplemental Table 1. The dependent variable was PSC discharge, which indicates whether an ADCG member was discharged before their service obligation was completed, analyzed as a dichotomous variable.

The values for these variables during the study period were defined as follows:

> -Most Common Primary Diagnosis Group: the most common primary diagnosis recorded on all outpatient visits for each unique patient.
>
> -Number of Visits: sum of all visits for each unique patient for each Most Common Primary Diagnosis Group.
>
> -Gender/Race Relative Risk: compares the proportion of members who sought BHC services, based on race and gender, to the proportion of ADCG who did not seek BHC services. Statistical Package for the Social Sciences (SPSS) 24 software was used to calculate the score for this variable.
>
> -Time in Service: the amount of time in an active-duty status each unique patient had at their first BHC appointment in 2016.

Missing data in the merged data set was treated as follows:

> -Values for *Time in Service* were imputed with the mean *Time in Service* according to the *Rank Group* to which they belonged.
>
> -Values for *Gender/Race Relative Risk* were (randomly) missing for males who did not have an associated race. They were imputed with the mean *Gender/Race Relative Risk* score for males of all races.

Cell sizes (n <10) were suppressed.

### Machine Learning Algorithm Testing and Selection

Training and test data sets were split between a 70:30 ratio^5^. To increase the statistical power of the data set and reduce the bias of the majority class (members who met their service obligation), the minority class was oversampled using the Synthetic Minority Over-sampling Technique (*SMOTE)* k nearest neighbor methodology^6^. The oversampled data set has 1,805 cases each of members discharged before completion of their obligated service and members who met their service obligation; in exploratory data analysis neither the merged data set model nor the oversampled merged data set were overfit (data not shown).

A combination of supervised learning and ensemble ML techniques^7^ were used for the analysis:

> -Decision Tree Classifier
>
> -Random Forest Classifier
>
> -Bagging Classifier
>
> -Gradient Boosting Classifier
>
> -Ada Boosting Classifier

Python™ programming language was used to apply algorithms for each ML model to the merged data set and oversampled merged data set. To screen the performance of each model, the following metrics were used:

> -Recall (sensitivity): true positives / (true positives + false negatives)
>
> -Precision: true positives / (true positives + false positives)
>
> -F1: 2*(Recall * Precision) / (Recall + Precision)

While these three metrics are interdependent, precision is the most important one by which to assess model performance, since it reveals how well the model predicts those who were discharged before completion of obligated service. Logistic regression, an established analytical epidemiologic approach^8^, was used as a methodologic comparator.

As appropriate, each of the models underwent hyperparameter tuning and feature engineering^9,10^. Because the *Number of Visits* variable had a significant number of outliers, values were log transformed to normalize the data. The *Age* variable was removed since it was highly correlated with *Time in Service* variable. The decision tree model’s complexity was reduced by limiting max tree depth to three, and samples per leaf to five. A random state of seven was used. K fold cross validation was used to validate the model for production using resampled data (every 10^th^ fold) from the training portion of oversampled merged data set. The grid search optimization function was used to find the best hyperparameters of the bagging classifier model. As a result, the model was optimized by limiting the base estimator max depth to three and max samples to 0.2. For the best-performing models with the highest precision, the ten leading features (predictive risk factors for early service termination) were examined using a matrix approach.

The Coast Guard Institutional Review Board reviewed this work and provided a non-research determination.

## Results

### Descriptive Analysis

We found that 26.4 of every 1,000 members who sought BHC services did not complete their service obligation within four years. Although females utilized more BHC services than males (p < .001), the proportion of males (2.7%) and females (2.4%) that were discharged before completion of their obligated service was not significantly different (p = 0.966). The *Race/Gender Relative Risk* variable did not reveal any statistically significant differences when comparing members discharge status (p = 0.56). The distribution of most common primary diagnosis group did not differ between members who met their service obligation and those who did not. Figure 1 demonstrates that both groups had the same three leading diagnosis groups (mood, anxiety, and adjustment disorders), and that the proportion of these diagnoses combined was similar (79 vs 83%).

**Figure 1:**
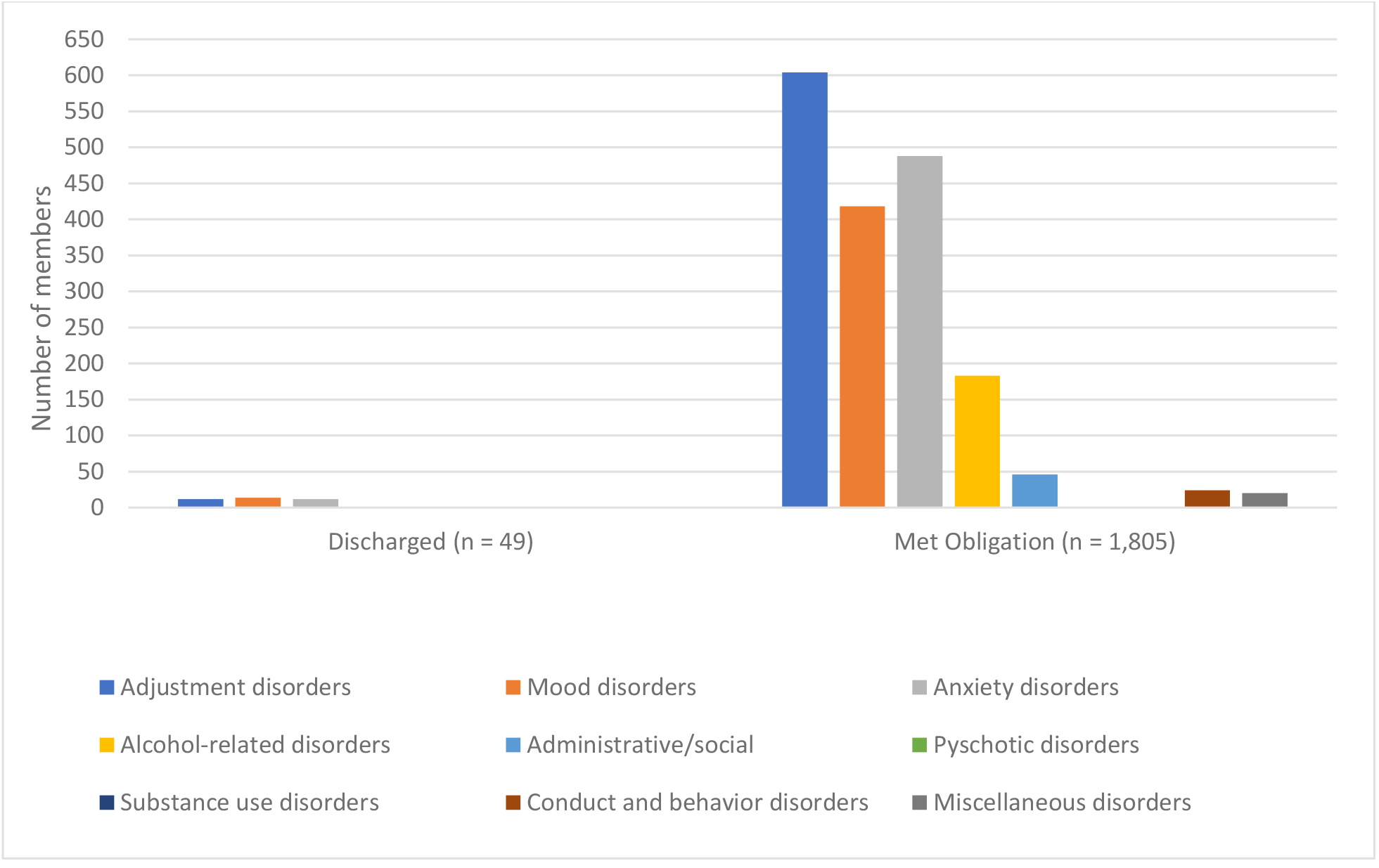
Number of Members Discharged Before Completion of Service Obligation and Members Who Met Obligation, by Most Common Primary Diagnosis Group

### Machine Learning Model Performance

The preliminary performance results for each of the ML algorithms are found in Table 1. The models on the oversampled data set provide the best performance results. Specifically, the precision scores for all models in the oversampled data set performed better than that of the merged data set. The three models with precision scores > 0.9 were bagging classifier, gradient boosting classifier, and logistic regression.

**Table 1.** Machine Learning Algorithm Performance Results, Oversampled Merged Data Set.

| Model | Recall | Precision | F1 Score |
| --- | --- | --- | --- |
| Logistic Regression | 0.837 | 0.901 | 0.901 |
| Decision Tree Classifier | 0.131 | 0.886 | 0.886 |
| Random Forest Classifier | 0.041 | 0.846 | 0.846 |
| Bagging Classifier | 0.049 | 0.963 | 0.963 |
| Gradient Boosting Classifier | 0.095 | 0.944 | 0.944 |
| Ada Boosting Classifier | 0.034 | 0.857 | 0.857 |

Following hyperparameter tuning and feature engineering, the models with the leading precision scores differed slightly, to include logistic regression, bagging classifier, and decision tree classifier (Table 2). Logistic regression was the best performing model (precision = 0.926), followed by the bagging classifier (0.911) and decision tree classifier (0.745).

**Table 2.** Final Results for Best Performing Algorithms Following Hyperparameter Tuning and Feature Engineering.

| Model | Recall | Precision | F1 Score |
| --- | --- | --- | --- |
| Logistic Regression | 0.893 | 0.926 | 0.910 |
| Bagging Classifier | 0.879 | 0.911 | 0.894 |
| Decision Tree Classifier | 0.875 | 0.745 | 0.805 |

Table 3 provides analysis of risk factors for early service termination for two of the three leading models, logistic regression and decision tree classifier; for methodological reasons features were not available for bagging classifier. Being White or Asian/Pacific Islander was a risk factor for early service termination in both logistic regression and decision tree classifier, as was senior enlisted rank (E-5 though E-9). The logistic regression model identified additional notable features predictive of discharge before completion of obligated service, including being male and having a primary diagnosis group of alcohol related disorders, notable since it differs from the three leading diagnosis groups seen in the descriptive results.

**Table 3.**
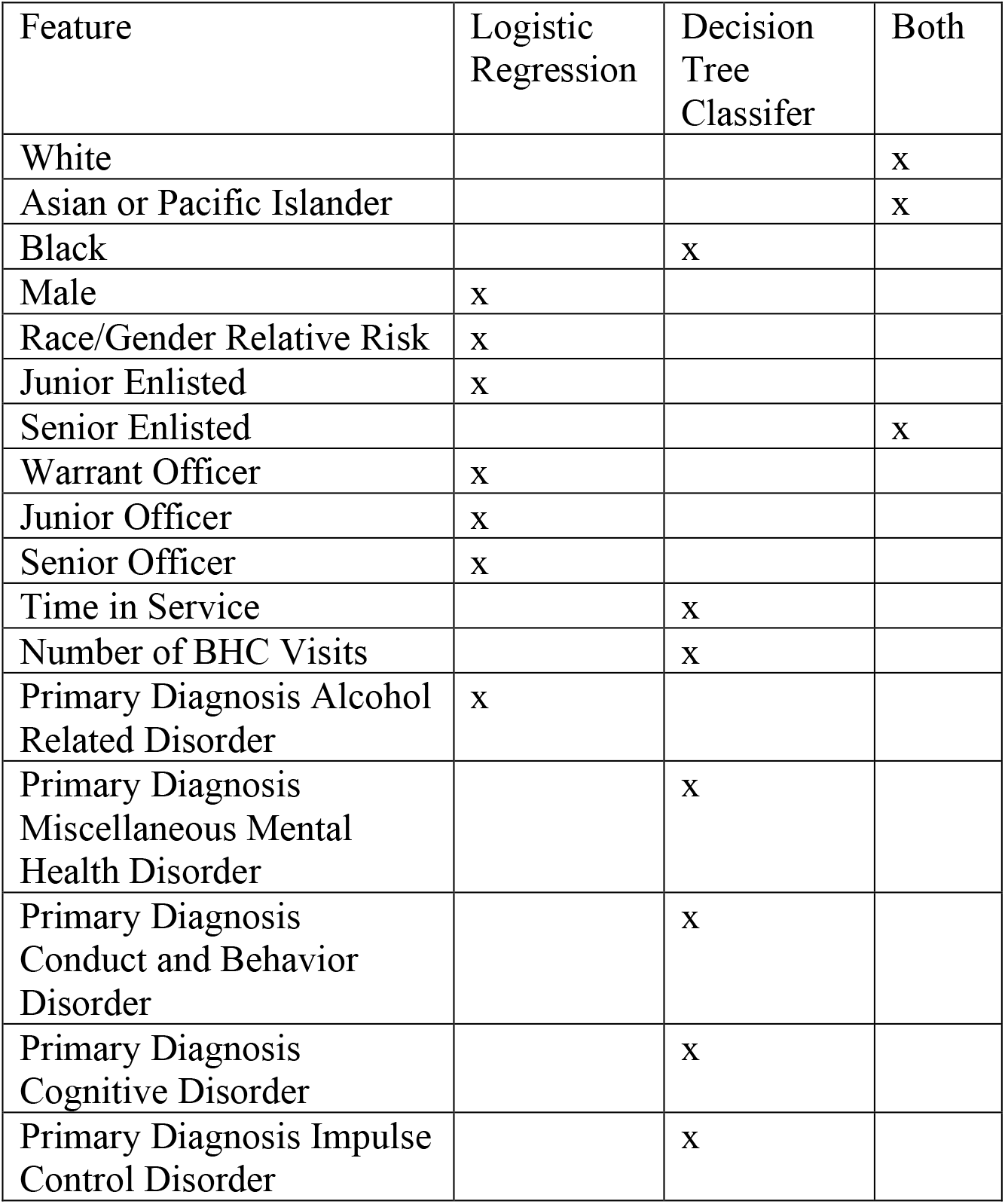
Top 10 Feature Matrix for Logistic Regression and Decision Tree Classifier Algorithms.

| Feature | Logistic Regression | Decision Tree Classifier | Both |
| --- | --- | --- | --- |
| White |  |  | x |
| Asian or Pacific Islander |  |  | x |
| Black |  | x |  |
| Male | x |  |  |
| Race/Gender Relative Risk | x |  |  |
| Junior Enlisted | x |  |  |
| Senior Enlisted |  |  | x |
| Warrant Officer | x |  |  |
| Junior Officer | x |  |  |
| Senior Officer | x |  |  |
| Time in Service |  | x |  |
| Number of BHC Visits |  | x |  |
| Primary Diagnosis Alcohol Related Disorder | x |  |  |
| Primary Diagnosis Miscellaneous Mental Health Disorder |  | x |  |
| Primary Diagnosis Conduct and Behavior Disorder |  | x |  |
| Primary Diagnosis Cognitive Disorder |  | x |  |
| Primary Diagnosis Impulse Control Disorder |  | x |  |

## Discussion

Although only 49 out of 1,854 cases in this study (2.6%) did not complete their obligated service, considering the operational, financial, and personal impacts of these discharges, it is worthwhile to learn more about the factors that may contribute to these losses. The high precision of the best-performing ML models provides a unique opportunity to understand factors that increase the probability a member will be discharged before completion of their service obligation. Recognizing the context, results, and limitations of this analysis can inform measures to improve service member retention.

The descriptive analysis yielded several notable results. The *Gender* variable produced mixed findings. While there was no difference between males and females in failure to complete service obligation, females in this study sought more BHC care. This may reflect longstanding differences in healthcare seeking behavior^11^. In the logistic regression model, male gender emerged as a leading feature for early service termination. This information may have implications for Coast Guard diversity, inclusion, and human resources policies and initiatives; for example, targeted efforts to encourage care-seeking among males can be considered.

The lack of difference in the diagnosis profile between members who did or did not complete their service obligation may affect BHC care provision and policy. This information should provide insight into the broad impact of mood, anxiety, and adjustment disorders on service members, and may provide the basis for a greater clinical focus on these common BHC conditions as a means to improve member retention. Similarly, the finding of alcohol use disorders as a risk factor for early termination in the logistic regression model identifies a condition amenable to intervention. Outpatient screening, brief intervention, and referral to care are both recommended and effective for such disorders^12^.

Among the shared risk factors for early termination seen in the logistic regression and decision tree classifier models, being in senior enlisted rank warrants comment. Possible explanations may include that seniority in the enlisted ranks introduces different technical, leadership, and managerial challenges. When mixed with the demands of assignment to an operational unit, the impact of BHC conditions could adversely affect member performance and well-being. The finding of Asian/Pacific Islander race/ethnicity as a shared risk factor is notable because of ongoing efforts to improve minority representation within the Coast Guard^13^. Several notes of caution are needed here: ML models applied in health care settings are potentially subject to racial and ethnic bias, and more broadly racial and ethnic disparities in healthcare and health outcomes are increasingly recognized as being due to structural racism^14,15^.

Although the mental and behavioral health diagnoses used in this analysis match those selected by the DHA AFHSD)^1^, we are not aware of other directly comparable analyses in other U.S. military services. While a prior analysis examined barriers to BHC as a predictor of intention to leave military service^16^, our study links receipt of BHC care to the outcome of early service termination in a longitudinal manner. Other previous studies have looked at attrition from service following specific events, such as deployments^17,18^ or particular BHC diagnoses^19^. These analyses, while not directly comparable, found much higher rates of attrition than we did in our examination.

This analysis is subject to at least three limitations. Service members seen for behavioral health care visits not captured in the M2 database were not included in this study. While the exclusion of care from non-mental health specialists limited the size of our sample, it may have strengthened our results, as obtaining mental health specialist care is generally considered a marker of greater illness severity.

The small number of members who were discharged before completion of obligated service (n = 49) created a moderate epidemiologic bias that was offset through oversampling. Another limitation was lack of access to case-level information surrounding the details of specific service member discharges, for example functional status or whether discharge was due to administrative separation or a medical board process. Strengths of this study include the four-year period of follow-up, and the use of multiple tuned ML models.

This analysis does not indicate there is lower retention among ADCG who seek outpatient BHC; nearly thirty nine out of forty ADCG who sought care completed service obligation. Thus, attrition from the Coast Guard in those who seek BHC is a rare outcome. By merging data sets this analysis evaluated key independent factors that had predictive validity for the outcome of early service termination. The final ML models, selected on the basis of feature engineering, provide the means to predict the probability a member will be discharged before completion of obligated service, as does logistic regression.

In this analysis, 26.4 of every 1,000 members who sought BHC services did not complete their service obligation within four years. This figure combined with the predictive power of the ML models has implications for the Coast Guard in relation to improving active-duty service member retention. In other settings such as higher education, machine learning has been useful to identify groups at risk for non-retention and target appropriate preventive interventions^20^. The Coast Guard has demonstrated an active interest in improving the mental and behavioral health of its service members^21^. Exit interviews or other qualitative data analysis (e.g., natural language processing of text from medical records) could complement these findings by clarifying the role of BHCs relative to other factors in decisions to terminate military service. Opportunities for future analyses across military services include identifying geographic trends, evaluating the impact of prevalent BHCs on service members in longer term studies, and determining career fields most affected by BHC diagnoses.

### Disclaimer

This work does not represent the official view of the United States Coast Guard, United States Department of Homeland Security, or the U.S. Public Health Service.

## Ethical guidelines statement

The U.S. Coast Guard Institutional Review Board reviewed this work and provided a non-research determination.

## Data availability statement

The source data is not publicly available.

