## Supplemental Table for "Service Retention Among Coast Guard Members Seeking Behavioral Healthcare"

Supplemental Table-Merged Data Set Independent and Dependent Variables

-Rank Group: Categorical variable.  This value groups military grades and ranks:

Cadet

>Enlisted, Junior (E1-E4)

>Enlisted, Senior (E5-E9)

>Officer, Junior (O1-O3)

>Officer, Senior (O4-O9, 10, 11)

>Warrant Officer (W1-W5)

-Most Common Primary Diagnosis Group: Categorical value.  The most common primary diagnosis group reported by M2 for all outpatient visits under the Mental Health Services Product Line by unique patients from 2015-2019. The diagnosis groups are classified by Agency for Healthcare Research and Quality (AHRQ) Clinical Classification System (CCS) and are derived from the ICD10 diagnosis codes.   The most common primary diagnosis groups reported for the population under study are:

>Adjustment disorders

>*Administrative/social admission

>Alcohol-related disorders

>Anxiety disorders

>Attention-deficit conduct and disruptive behavior disorders

>Delirium dementia and amnestic and other cognitive disorders

>Impulse control disorders

>Miscellaneous mental health disorders

>Mood disorders

>Personality disorders

>**Residual codes; unclassified

>Schizophrenia and other psychotic disorders

>Substance-related disorders

*[drug disorders](https://safe.menlosecurity.com/doc/docview/viewer/docN10C02E656A7F9ae2734c288801069a731eb6f70640b2c074c170f6d2108ed9fa1be59269f84b)

**other mental health disorders not elsewhere classified

-Race: Categorical variable. The race categories included in the analysis are:

White

>Hispanic

>American Indian/Alaskan Native

>Asian or Pacific Islander

>Black

>American Indian/Alaskan Native

>***Unknown

***2% of race categories that did not show agreement or were missing between the M2 data set and PSC data sets were classified as Unknown.

-Gender: Categorical variable. The biological sex of the patient:

>Male

>Female

-Unit Type: Categorical variable. The type of Coast Guard operational unit the patient was assigned to at their first outpatient visit in 2016.  Unit types were divided into three categories:

>Cutter

>Station

>Air Station

-Number of Visits: Discrete variable. The most common primary diagnosis group that had the most outpatient mental and/or behavioral health visits as reported by M2 under the Mental Health Services Product Line by unique patients see in 2016 from 2016-2019.

-Gender/Race Relative Risk: Continuous variable. Measure of the ratio of the probability that an AD CGSM, based on race and gender, sought MH services to the probability that those AD CGSMs, based on race and gender, did not seek MH services. A Gender/Race Relative Risk score was assigned to each the following categories:

>American Indian/Alaskan Native Female

>American Indian/Alaskan Native Male

>Black Female

>Black Male

>Asian or Pacific Islander Female

>Asian or Pacific Islander Male

>Hispanic Female

>Hispanic Male

>Unknown Race Female

>Unknown Race Male

>White Female

>White Male

-Time in Service: Continuous variable. Time on active duty at their first MH appointment during calendar year 2016.

Dependent variable:

-PSC Discharge Reason: Categorical variable (binary).   The following discharge types or AD status are classified according to whether the AD CGSMs completed their service obligation:

Discharge types or other that indicate completion of service obligation or continued service through 2019 (referred to as “members who met their service obligation” or similar throughout paper):

>Remain on AD 2019

>Retirement

>Enlisted Separation Request

>Voluntary Separation

>Sufficient service for retirement

>Completion of required active service

Discharge types that indicate incomplete service obligation (referred to as “members discharged before completion of service obligation” or similar throughout paper):

>Mandatory Separation

>Misconduct

>Fraudulent Entry into Military Service

>Placed on Permanent Disability Retirement List

>Adjustment Disorder

>Separation for Miscellaneous/General Reasons

>Unsatisfactory Performance

>Unacceptable Conduct

>Maximum service or time in

>Disability, permanent

>Condition, not a disability
